# Prevalence of Chronic Rhinosinusitis in Rajavithi Hospital Workers, Bangkok, Thailand

**DOI:** 10.1101/2023.07.20.23292827

**Authors:** Wirach Chitsuthipakorn, Suphitsara Naphapunsakul, Patlada Korwattanamongkol, Kittichai Mongkolkul, Somjin Chindavijak

**Affiliations:** College of Medicine, Rangsit University, Bangkok, Thailand; Otolaryngology department, Rajavithi hospital, Bangkok, Thailand

**Keywords:** Prevalence, Chronic rhinosinusitis, nasal endoscopy, Rajavithi hospital, Bangkok

## Abstract

**Background:** The prevalence of chronic rhinosinusitis (CRS) has been reported to range from 5% to 12% in different parts of the world. However, the prevalence of CRS in Thailand has not been investigated. Our objective was to determine the prevalence of CRS among hospital workers and identify any potential problems encountered during the survey.

**Methods:** Adult workers (>18 years) of Rajavithi Hospital, a tertiary hospital in Bangkok, Thailand, were recruited. This cross-sectional survey was conducted from September 2021 to September 2022. The participants were given a link to online questionnaires asking if they had nasal obstruction, discharge, decreased smell sensation, or facial pain and their respective duration. Phone numbers and email were asked for contact when needed. The participants who fulfilled the symptom and duration according to the European position paper on rhinosinusitis and nasal polyp 2020 were counted as symptom-based CRS patients. These patients were then contacted for nasal endoscopy. The endoscopic-based diagnosis of CRS was made after positive endoscopy findings.

**Result:** A total of 1,025 participants (mean age of 33.7 years) were recruited. Of total, there were 34 (3.3%) and 6 (0.58%) participants fulfilled symptom- and endoscopy-based diagnoses, respectively. Fourteen participants did not respond to calls or emails. Five patients refused to visit the clinic due to inconveniences. One patient refused nasal endoscopy because of the expenses.

**Conclusion:** The overall prevalence of chronic rhinosinusitis in workers of Rajavithi Hospital, was 3.3% and 0.58% by symptom- and endoscopy-based criteria, respectively. Difficulty reaching the participants for nasal endoscopy was the leading survey problem.

## INTRODUCTION

The prevalence of chronic rhinosinusitis (CRS) has been reported in various geographical locations, ranging from 5% to 12% in different populations.^1^ A European study reported the mean prevalence of CRS at 10.9%, with a range of 6.9% to 27.1% based on data from 12 countries.^2^ Prevalence studies in other parts of the world found rates of 5.51% in Sao Paulo,^3^ 6.8% in China,^4^ 6.95% in Korea,^5^ and 12% in the United States.^6^

CRS is a chronic disease that requires both surgical and long-term medical management. Therefore, prevalence of the disease may determine the magnitude of the problem in health economics and serves as a basis for resource allocation and medical services. It draws attention from policymakers to assess the cost-effectiveness of current and novel treatment like biologics. The European position paper on rhinosinusitis and nasal polyp 2020 (EPOS2020) has defined two diagnostic criteria for CRS in epidemiological studies.^2^ First, the symptom-based or epidemiological diagnosis relies on symptoms and duration. Second, the endoscopy-based or clinical diagnosis criteria require the presence of symptoms along with objective evidence.

The prevalence of CRS in Thai adults has not yet been determined. Conducting a nationwide population-based survey, which may be time-consuming and require a considerable budget, is necessary. Therefore, we conducted this pilot study among workers at Rajavithi Hospital in Bangkok, Thailand, to assess the prevalence of CRS using both symptom-based and endoscopy-based criteria and to identify any problems that may arise during the survey before conducting a larger study.

## METHOD

### Participants

The study protocol received approval from the institutional review board of Rajavithi Hospital, Bangkok, Thailand, on 27^th^ August 2021 (no. 208/2564) and was registered in Thai Clinical Trial Registry number TCTR20210922007, on 22^nd^ September 2021. The cross-sectional survey was conducted from September 2021 to September 2022. Participants were recruited through verbal communication by all authors, hospital circulating letters, or social network communications. Informed consent was obtained from participants through voluntary scanning of the QR code or clicking the online link provided either verbally or in the letter. Participants had to be at least 18 years old and have a good understanding of the Thai language in both reading and speaking.

### Instruments and data collection

The demographic data, including gender, age, occupation, and residential district, were collected. Other factors such as influenza vaccination, smoking status, stress level, underlying diseases, and education level were also recorded. Participants were asked to provide their email addresses and/or phone numbers for contact purposes, based on their comfort level.

The four symptoms assessed were nasal congestion, nasal discharge, impaired sense of smell, and facial pain, along with their respective durations. According to EPOS2020 guidelines, CRS criteria were defined as a combination of at least two out of the four mentioned symptoms, with at least one symptom (either nasal obstruction or nasal discharge) persisting for a minimum of 12 weeks.^2^ Participants whose symptoms and duration matched the CRS criteria were identified as symptom-based CRS patients. Participants who met the symptom-based criteria were then contacted for a formal nasal endoscopic assessment at the Rajavithi hospital ENT clinic. The clinical diagnosis, based on symptoms and positive nasal endoscopy findings, was made by the rhinology staff (W.C.), Table 1.

**Table 1.** Definitions of CRS according to the European Position Paper on Rhinosinusitis and Nasal Polyp 2020.

|  |
| --- |
| Definition of CRS: Inflammation of the nose and the paranasal sinuses characterized by two or more symptoms, one of which should be either nasal blockage/obstruction/congestion or nasal discharge (anterior/posterior nasal drip) |
| **Symptom-based CRS**<br>Nasal blockage/obstruction/congestion or nasal discharge (anterior/posterior nasal drip) ± Facial pain/pressure, ± loss of smell.<br>Duration of symptoms > 12 weeks |
| **Endoscopy-based CRS**<br>Symptom-based CRS <u>AND</u> Endoscopic finding: the presence of polyps, presence of edema in the middle meatus or presence of thick purulent discharge in the middle meatus to define endoscopy positive CRS. |

Abbreviations: CRS=chronic rhinosinusitis

### Outcomes

The main outcome of this study was to determine the prevalence of CRS among workers at Rajavithi Hospital by considering both symptom-based and endoscopy-based criteria. Additionally, the study aimed to identify any potential challenges or issues that may occur during the survey process in order to address them before conducting a larger-scale study.

### Statistical analyses

Descriptive statistics were used to summarize the demographic characteristics of the participants, including gender, age, occupation, and residential district. The prevalence of CRS was calculated by dividing the number of participants who met the CRS criteria by the total number of participants in the study. The prevalence rate was expressed as a percentage with a corresponding 95% confidence interval (95%CI). To assess the association between CRS and various factors such as influenza vaccination, smoking, stress level, underlying disease, and education, appropriate statistical tests were performed. Chi-square test or Fisher’s exact test was used for categorical variables, while t-test or Mann-Whitney U test was used for continuous variables, depending on the distribution of the data. All statistical analyses were performed using statistical software Stata 17 at a significance level of p < 0.05.

## RESULTS

A total of 1,025 participants with a mean age of 33.7 were enrolled in the study, consisting of 829 (80.9%) females and 196 (19.1%) males. The age distribution showed that 767 (74.8%) participants were between 18 and 40 years old, 241 (23.5%) were between 41 and 60 years old, and 17 (1.65%) were above 61 years old. Regarding smoking status, 26 (2.50%) participants reported being smokers, while 999 (97.5%) reported not smoking.

Among the total participants, 34 (3.3%) met the criteria for symptom-based diagnosis of CRS. However, out of these 34 participants, 20 were not evaluated with nasal endoscopy. Among the remaining 14 participants who underwent nasal endoscopy, 6 had positive pus discharge from the sinus ostium, confirming the diagnosis of CRS based on endoscopy findings. Thus, the prevalence of CRS based on endoscopy-based diagnosis was 0.58% of the total study population. Table 2 provides a summary of the demographic of participants and numbers of CRS patients in each category.

**Table 2.**
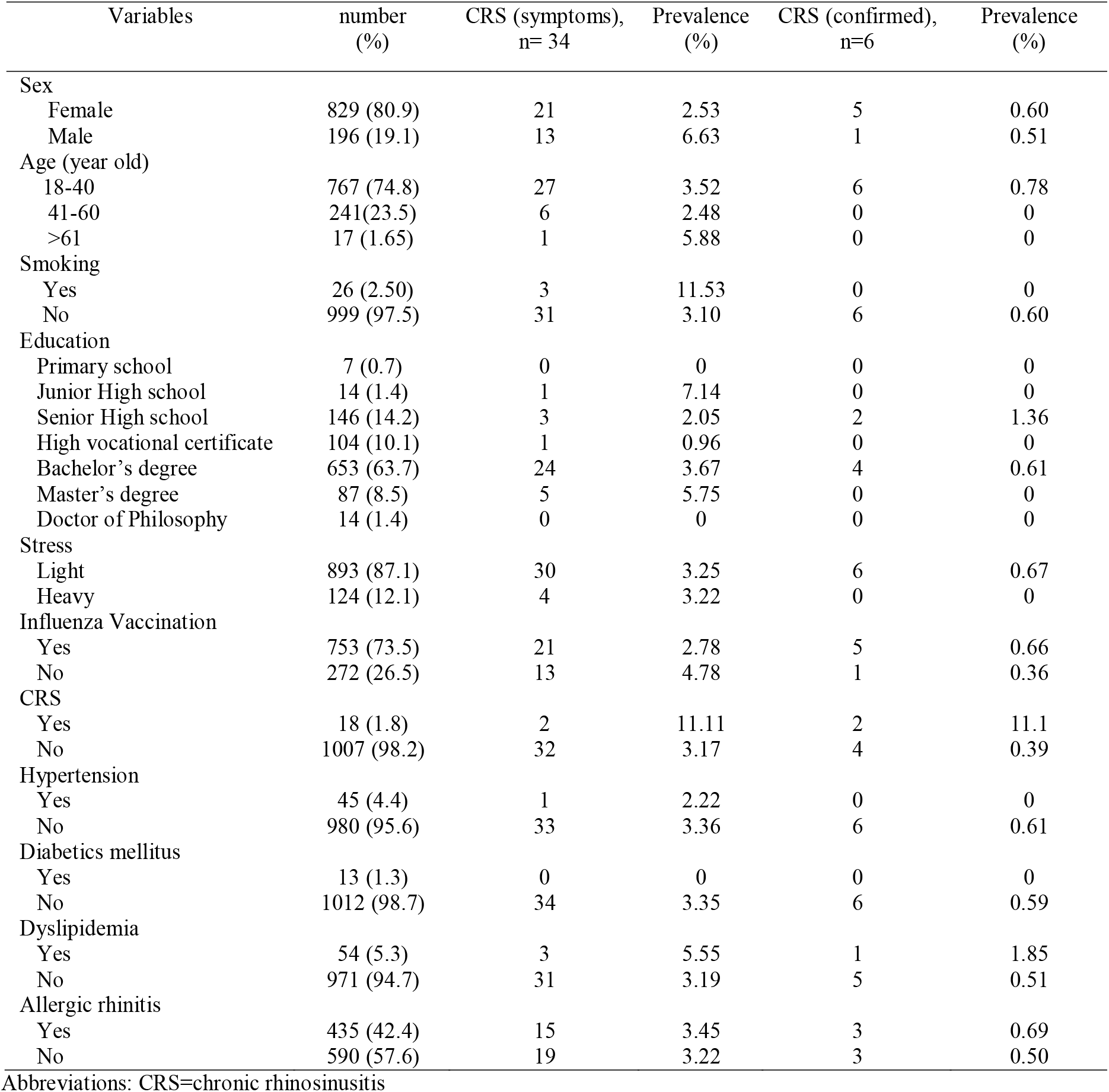
Demographics of included participants and number of CRS patients with respect to each characteristic.

| Variables | number<br>(%) | CRS (symptoms),<br>n= 34 | Prevalence<br>(%) | CRS (confirmed),<br>n=6 | Prevalence<br>(%) |
| --- | --- | --- | --- | --- | --- |
| Sex |  |  |  |  |  |
| Female | 829 (80.9) | 21 | 2.53 | 5 | 0.60 |
| Male | 196 (19.1) | 13 | 6.63 | 1 | 0.51 |
| Age (year old) |  |  |  |  |  |
| 18-40 | 767 (74.8) | 27 | 3.52 | 6 | 0.78 |
| 41-60 | 241(23.5) | 6 | 2.48 | 0 | 0 |
| >61 | 17 (1.65) | 1 | 5.88 | 0 | 0 |
| Smoking |  |  |  |  |  |
| Yes | 26 (2.50) | 3 | 11.53 | 0 | 0 |
| No | 999 (97.5) | 31 | 3.10 | 6 | 0.60 |
| Education |  |  |  |  |  |
| Primary school | 7 (0.7) | 0 | 0 | 0 | 0 |
| Junior High school | 14 (1.4) | 1 | 7.14 | 0 | 0 |
| Senior High school | 146 (14.2) | 3 | 2.05 | 2 | 1.36 |
| High vocational certificate | 104 (10.1) | 1 | 0.96 | 0 | 0 |
| Bachelor's degree | 653 (63.7) | 24 | 3.67 | 4 | 0.61 |
| Master's degree | 87 (8.5) | 5 | 5.75 | 0 | 0 |
| Doctor of Philosophy | 14 (1.4) | 0 | 0 | 0 | 0 |
| Stress |  |  |  |  |  |
| Light | 893 (87.1) | 30 | 3.25 | 6 | 0.67 |
| Heavy | 124 (12.1) | 4 | 3.22 | 0 | 0 |
| Influenza Vaccination |  |  |  |  |  |
| Yes | 753 (73.5) | 21 | 2.78 | 5 | 0.66 |
| No | 272 (26.5) | 13 | 4.78 | 1 | 0.36 |
| CRS |  |  |  |  |  |
| Yes | 18 (1.8) | 2 | 11.11 | 2 | 11.1 |
| No | 1007 (98.2) | 32 | 3.17 | 4 | 0.39 |
| Hypertension |  |  |  |  |  |
| Yes | 45 (4.4) | 1 | 2.22 | 0 | 0 |
| No | 980 (95.6) | 33 | 3.36 | 6 | 0.61 |
| Diabetics mellitus |  |  |  |  |  |
| Yes | 13 (1.3) | 0 | 0 | 0 | 0 |
| No | 1012 (98.7) | 34 | 3.35 | 6 | 0.59 |
| Dyslipidemia |  |  |  |  |  |
| Yes | 54 (5.3) | 3 | 5.55 | 1 | 1.85 |
| No | 971 (94.7) | 31 | 3.19 | 5 | 0.51 |
| Allergic rhinitis |  |  |  |  |  |
| Yes | 435 (42.4) | 15 | 3.45 | 3 | 0.69 |
| No | 590 (57.6) | 19 | 3.22 | 3 | 0.50 |

Abbreviations: CRS=chronic rhinosinusitis

Among the 20 participants who were not evaluated, 14 did not respond to calls, emails, or both, 5 refused to visit the clinic due to conflicts with clinic operating times and their working schedules, and 1 patient declined nasal endoscopy due to concerns about the associated expenses.

Over a period of four months, which accounted for 33% of the total study duration, non-direct communication methods such as messaging via social groups or circulating letters between hospital departments were primarily used. A total of 199 participants, representing 19.4% of the total participants, were successfully enrolled through these methods. The remaining 80.6% of enrollments were completed during the subsequent eight months, comprising 66% of the study duration. This phase involved a combination of both direct and indirect communication approaches with hospital workers to enhance participation. Of all participants, 352 (34.3%) did not provide their phone numbers in the online survey, despite explicitly provided privacy statement.

## DISCUSSION

This pilot study aimed to assess the prevalence of chronic rhinosinusitis (CRS) among workers at Rajavithi Hospital in Bangkok, Thailand. The findings revealed a prevalence of 3.31% for symptom-based CRS and 0.58% for endoscopy-based CRS in the study population. These figures are lower than the prevalence rates reported in other countries, which typically range from 5.5% to 12%.^1,7–9^

Among the participants who qualified for symptom-based CRS, only 6 individuals underwent nasal endoscopy and were confirmed as having CRS, resulting in a prevalence of 0.58% for endoscopy-based CRS. However, if all 20 participants who qualified for symptom-based CRS had undergone nasal endoscopy and were positive, the prevalence could have been as high as 2.53%. This suggests that the true prevalence of endoscopy-based CRS in the study population could range between 0.58% and 2.53%, which is comparable to the reported prevalence of 1.01-1.2% in South Korea using similar criteria.^5,10^

The low response rate from messaging or circulating letters during the initial phase of the survey highlights the limitations of relying solely on non-direct communication methods. Face-to-face enrollment not only expedited the recruitment process but also built trust and provided an opportunity to address any questions or concerns. Despite explicitly providing a privacy statement, 34.3% of the total participants did not provide their phone numbers in the online survey. This could suggest concerns about privacy or other reasons still exists. The lack of phone numbers provided by participants hindered effective communication, potentially leading to an underestimation of the prevalence. In future larger-scale studies, the inclusion of mandatory phone number fields or a clear direct communication about the purpose of collecting this information may help improve input rates.

Five participants (15%) did not comply with clinic visits for nasal endoscopy. Our clinic operates during daytime opening hours (9am - 4pm) of the hospital, which is likely overlapping to the daytime shift of other hospital workers. To avoid this, we could prepare 3-4 portable nasal endoscope sets for 100 enrollments in the future study and be ready for the procedure when a symptoms-based CRS candidate appears.

It is important to acknowledge the limitations of this survey. The study participants only represented a specific group of individuals working in the hospital, which may introduce selection bias and limit the generalizability of the findings to the broader population. Additionally, the skewed distribution of age and sex in the participant characteristics may further affect the generalizability of the results. Post hoc risk factor analysis was not performed in this pilot study due to the heavily skewed distribution and was not the intended purpose of this study. However, identifying potential risk factors for CRS would be valuable in understanding the disease better and developing appropriate preventive strategies.

Despite these limitations, this study provides an initial benchmark for CRS prevalence among small group of Thai people. Lessons learned from the challenges faced during this study will inform the planning and execution of larger-scale surveys in the future, including proper sampling processes, enhanced participant engagement, and improved data collection. A national-level study with a more representative sample would be essential to accurately determine the magnitude of the CRS problem in Thailand.

## CONCLUSION

The overall prevalence of chronic rhinosinusitis at Rajavithi Hospital in Bangkok was 3.3% based on symptoms and 0.58% based on endoscopy criteria, respectively. Direct communications should be encouraged to improve enrolments.

## Data Availability

All data produced in the present study are available upon reasonable request to the authors

## ACKNOWLEDGEMENT

None

